# Exploring the views of people living with pulmonary fibrosis and health professionals on genetic testing in PF: A qualitative study

**DOI:** 10.64898/2026.08.03.26359293

**Authors:** Sarah Rawlings, Narelle S Cox, Ching Shan Wan, Joanne Dickinson, Anne E Holland

**Author notes:** **Corresponding author** Mrs Sarah Rawlings Respiratory Research@Alfred Monash University, Level 6, 99 Commercial Road,Melbourne VIC 3004, Australia.

## Abstract

**Objectives:** Genetic testing is increasingly used in the diagnosis and management of respiratory conditions, including pulmonary fibrosis (PF). The perspectives of people with PF and healthcare professionals (HCP) on the use of genetic testing remain largely unexplored.

**Methods:** A qualitative study was undertaken. People living with PF, their caregivers, and HCP were invited to undertake a semi-structured interview. Interviews were conducted via videoconference or telephone, audio-recorded, and transcribed verbatim. Data were analysed by two researchers using inductive thematic analysis.

**Results:** Thirty-eight participants; 15 people living with PF, 1 caregiver, and 22 HCPs were interviewed. Analysis revealed three key themes. Genetic testing in PF was valued by all groups; people with PF wanted testing now, whilst respiratory physicians were cautious, citing their uncertainty regarding clinical value. All groups desired more information and support; people with PF desired a better understanding of terminology, whilst genetic counsellors wanted to better understand PF. No single model for returning genetic results in PF was identified, however resources, multidisciplinary care, and timely return of results was considered important.

**Conclusion:** Genetic testing is valued by people with PF and their HCP, but uncertainties remain regarding whether it should be offered and how results should be best communicated.

## Introduction

Pulmonary Fibrosis (PF) is a complex and rare disease, characterised by cough, shortness of breath, and fatigue(1). Symptoms are broad and non-specific, often overlapping with other respiratory conditions, contributing to lengthy delays in diagnosis or misdiagnosis(2). In the absence of a single definitive diagnostic test for PF, accurate diagnosis relies on integrating clinical, radiological, and pathological findings, supported by multidisciplinary discussion(3). With the growing appreciation of the utility of genomics in healthcare delivery(4, 5), genetic testing is increasingly recognised as a useful supplementary tool to enhance diagnostic certainty in PF(6). Genetic testing provides useful insights into familial risk, disease progression, and can inform management of disease(7, 8). However, prevalence of testing remains low(6, 7), and practise is variable(6).

Little is known about how people with PF and relevant stakeholders, including geneticists, genetic counsellors, and respiratory physicians, perceive genetic testing for PF. In this qualitative study we aimed to understand the perspective and preferences of people living with PF and their caregivers and health professionals (HCP) on genetic testing including disclosure of results.

## 2. Methods

### Study design and participants

A qualitative study was conducted between November 2024- February 2025. Approval was obtained from Monash University Human Research Ethics Committee (Project ID: 42685). All participants provided written informed consent.

People living with PF, their caregivers and HCPs were invited to participate in a one off semi-structured interview. People living with PF were invited to participate through the PF Australasian Clinical Trial Network e-newsletter, using convenience sampling. Eligible individuals had PF or cared for someone with PF. Healthcare professionals were invited to participate through study promotion in the Thoracic Society of Australia and New Zealand, Human Genetics Society of Australasia (HGSA), and the Australasian Society of Genetic Counsellors e-newsletters. Combined sampling strategies, purposeful and snowball, were employed to enhance diversity of HCP working in non-tertiary centres, who are difficult to reach(9). Healthcare professionals were eligible irrespective of prior experience with genetic testing. Participants were excluded if they could not provide informed consent. Reporting followed the Consolidated Criteria for Reporting Qualitative Research (COREQ).

Demographic data including age, gender, residential location (metropolitan or regional), and prior experience with genetic testing, was collected for people living with PF. Gender, profession, workplace location (categorised as metropolitan or regional), and experience with genetic testing was recorded for HCPs.

### Procedure

Semi-structured interviews were conducted exploring participant perspectives of genetic testing in PF. Interviews were completed via videoconferencing (Zoom) or telephone. An interview guide was designed, and pilot tested with a local community advisory member, who provided feedback on the clarity and sequencing of questions (Supplementary material, Table S1 and S2). Interviews were conducted by a researcher (SR) with experience in qualitative research methodology who was not involved in the care of patients. Interviews were audio recorded and transcribed verbatim. Field notes were taken during the interview process. Data collection was completed until no new themes were observed(10).

### Data analysis

Interview transcripts were analysed by two researchers independently (SR and CSW), both experienced with qualitative data analysis. Data coding was stored and organised using NVivo(11). Inductive thematic analysis was conducted, using a six-step approach(12). Researchers individually familiarised themselves with the data, completed line-by-line analysis, and identified descriptive codes from the data. Related codes were collapsed creating major themes and subthemes, which were reviewed and refined. Researchers met to discuss themes and subthemes until consensus was reached. Reflexivity was practiced throughout the process. Relevant quotations were extracted, providing supportive data for each theme. Data collection and analysis were conducted concurrently, allowing emerging themes to guide interviews until data saturation was reached and no new information arose(10, 13).

## 3. Results

A total of thirty-eight participants were interviewed (15 people living with PF, 1 caregiver, 22 HCPs). Demographics of people living with PF and HCPs can be found in Tables 1 and 2 respectively.

**Table 1.** Demographics of people with lived experience of PF.

|  | People living with PF<br>N=15 | Carer<br>N=1 |
| --- | --- | --- |
| Gender, F | 7 (47%) | 0 (0%) |
| Age range, yrs. |  |  |
| Under 60 | 1 (7%) | 0 (0%) |
| 60 and over | 14 (93%) | 1 (100%) |
| Prior experience with testing, N | 13 (87%) | 1 (100%) |

Abbreviations F: female; yrs: years; N: No prior experience with testing

**Table 2.** Demographics of health care professionals.

| Profession | Respiratory<br>Physician<br>N=13 | Genetic<br>Counsellor<br>N=2 | Geneticist<br>N=4 | Nurse<br>N=2 | Other<br>profession<br>N=1 |
| --- | --- | --- | --- | --- | --- |
| Gender, F | 4 (31%) | 2 (100%) | 2 (50%) | 2 (100%) | 1 (100%) |
| Workplace location |  |  |  |  |  |
| Metro, % | 11 (85%) | 2 (100%) | 4 (100%) | 2 (100%) | 1 (100%) |
| Regional, % | 2 (15%) | NA | NA | NA | NA |
| Prior experience with testing,<br>N | 5 (38%) | 0 (0%) | 0 (0%) | 0 (0%) | 0 (0%) |

Abbreviations F: female; N: No prior experience with testing

### People living with PF

People living with PF were aged between 57 and 92 years, 47% were female (Table 1). Nine (56%) participants lived in metropolitan locations, while six (44%) lived regionally. People living with PF were diagnosed between 2 years and 14 years prior to enrolment, and two people had personal experience with genetic testing as part of their PF diagnosis. The median [IQR] interview duration was 49.5 [39.5 to 53] minutes.

### Health professionals

Twenty-two HCPs were interviewed, including respiratory physicians, a haematologist, ILD nurses, clinical geneticists, and genetic counsellors (Table 2). Five (38%) respiratory physicians had no prior genetic testing experience, 2 (15%) had some experience, and 6 (46%) were experienced. Median [IQR] clinician interview duration was 29.5[25 to 33] minutes.

Three overarching themes on the perspective of genetic testing in PF emerged. Within each theme, subthemes illustrate similarities and differences in participant viewpoints (Supplementary Tables S3- S5).

### Theme 1: Genetic testing is valued

#### People with PF and HCP recognise the value of genetic testing, but ongoing barriers exist

Genetic testing was valued across all groups, with a shared recognition of its emerging clinical relevance and future role to support PF diagnostic clarity. People living with PF valued testing to understand inheritance and familial links.

> *‘I’m wondering whether there’s any family relationship. I certainly would like to know for my sister’s benefit as well’(P12)*.Health professionals across all disciplines viewed genetic testing as a promising tool, supporting predictive modelling and precision medicine. Respiratory physicians expressed interest in genetic testing, viewing its importance to *‘add to the story about what the chances were of developing progressive disease’(RP_9).* All HCPs reported systemic barriers to the uptake of genetic testing in PF, including poor access to testing services, workforce constraints such as limited availability of geneticist and genetic counsellors, an absence of standardised pathways, and inadequate funding. Genetic counsellors expressed concern around the limited resources directed to familial follow up. Meanwhile, respiratory physicians who had not previously ordered genetic tests acknowledged their lack of clinical genetics expertise (Table S3).

### People with PF are ready to embrace testing now

People living with PF described having the option of genetic testing to be *‘an absolute godsend’(P19).* They reported it was *‘great that it’s (genetic testing) being given attention’(P18)*, with the majority stating they were ready to embrace testing now (Table S3).

People living with PF reported to be driven towards genetic testing to understand their disease in more detail, describing additional disease knowledge as *‘an added bonus’(P14)*.

> *‘The more information the better because its information that can assist you to deal with it’(P18)*.Despite the infancy of genetic testing, people living with PF overwhelmingly reported that testing renewed their hope for improved disease treatments and outcomes, even if not their own.
>
> *‘It’s really not going to change my outcome, but it could potentially change other people’s outcomes’(P8)*.

***Respiratory physicians remain hesitant, driven by uncertain implications for clinical practise.*** Respiratory physicians without prior genetic testing experience report their hesitancy was driven by evolving knowledge of the genetics of PF. These HCPs identified that they would require greater knowledge and evidence of the clinical utility before testing was embedded into routine care (Table S3).

*When we order a test, we’ve got to have an idea about how we’re going to act on these results. Until I understand what the test means then I wouldn’t really be comfortable in ordering it’(RP_12)*.

Respiratory physicians currently utilising genetic testing describe uncertainty surrounding variants of uncertain significance, how to interpret their clinical significance and actionability of results.

> *‘(Variants of uncertain significance) That’s part of the reason why we don’t do routine genetic testing. Because we don’t really know what to do with them or what their significance is. And there’s no difference in how you treat the patients for it’’ (RP_1)*.

***Theme 2: More information and support are desired, but needs differ***.

### Genetic knowledge is variable across people living with PF and respiratory physicians, other professionals lack PF disease knowledge

People with PF who had undergone genetic testing reported that information provided to them on testing was overwhelming, stating ‘*it was beyond my scope of understanding. Current resources are not fit for purpose’ (P18)*. Similarly, genetic counsellors reported that many people with PF referred for their support had a poor understanding of genetic testing principles (Table S4).

Physicians with no testing experience reported limited knowledge on basic testing process; how to access testing and what test to use. Physicians who had prior experience reported to have adequate knowledge around testing pathways, however lacked knowledge and confidence around how to interpret genetic results. Genetic counsellors reported their uncertainty centred around PF specific disease knowledge and how to apply genetic findings meaningfully into clinical care (Table S4).

> *‘I don’t have all the clinical knowledge and a lot of the questions and things that come up from patients is the clinical side of things because that’s what they’re dealing with’(GC_2)*.

### People with PF want resources and accessible support to understand genetic language

Most people living with PF report that they need clear resources explaining the objectives of genetic testing, the steps involved, and how results are returned, *‘giving clarity of the (testing) procedure’(P19).* Some people had a preference toward written resources *‘something I can study and try and understand’(P12)*, while others desired the use of online material including video resources.

Some participants suggested seeking ‘*information from other patients who have experienced genetic testing’(P12)*, as a helpful way to clarify testing processes.

People living with PF additionally reported having adequate time to comprehend genetic information was important, as was having accessible support from HCPs, including someone who could be easily reached to ask a question (Table S4).

### Health professionals want training, pathways, and clinical support

Respiratory physicians, regardless of testing experience, reported a need for genetic testing protocols and education on how to refer for testing, how to carry out testing including *‘where to send blood’ (RP_9)* and how to disclose results. Health professionals suggested a variety of resource modalities; some preferred online resources, others face to face seminars or peer learning to support knowledge needs. However, despite the benefits of working as a team, health professional still desired ‘*resources’(GC_1*) (Table S4).

### Theme 3: No consensus on the optimal model for returning genetic results in PF

All groups emphasised the importance of receiving genetic information as soon as possible. Views were varied on who was best placed to disclose results. Some people living with PF reported no strong preference around who should return their genetic results, while others preferred a specialist, and others still preferred a clinician they already knew (Table S5).

> *‘Anyone, to tell you the truth. I wouldn’t be fussy. Whatever would be the most convenient way for you.’ (P8)*
>
> *‘Probably my GP I have known for a very long time or someone that knows all about it. So that you’ve got that bit of knowledge behind so that if there’s questions, they can be answered.’(P13)*Respiratory physicians who had experience with reported results suggested it *‘should be always delivered by a clinician expert in those decisions’(RP_2).* Geneticists and genetic counsellors reported that ‘*the person that orders the test should give the results’(*Gen_4). Genetic counsellors also reported respiratory physicians played an important part in the return of results as they had the clinical knowledge about PF. However, acknowledged that physicians may not *‘have the time nor necessarily the understanding of all the different pathways of how to deal with all of the implications that come with doing genetic testing’(*GC_2), particularly the skills to support familial testing and follow up. Overwhelmingly, all HCPs agreed the return of PF genetic results was not yet suitable for primary care.

### Logistics to support return of genetic result includes resources and multidisciplinary support and collaboration

People living with PF suggested logistics to support the return of genetic results, including the provision of handouts *‘something they could read themselves’(P14)*. Some participants stated an acceptance of electronic communication, while others preferred face-to-face explanations. Regardless, appropriate discussion was important, *‘I am happy for the information to come via email, but it’s always helpful, if there are any sort of out of whack results, to follow that up with a discussion with a medico of some kind’(P1).* Ultimately, people living with PF reported there was no *‘one size fits all approach’(P4)* to return of results, however multidisciplinary collaboration between healthcare providers gave them reassurance, trust, and a sense of comfort *‘knowing it was not just one person*’ making all the decisions (Table S5).

Health professionals across disciplines reported similar views, with no single preferred model for returning results. All HCPs reported collaborative multidisciplinary support and resources would support result disclosure. Physicians reported professional collaboration with clinical geneticists and genetic counsellors enhanced their knowledge and confidence in genetic testing practices. Genetic counsellors valued input with respiratory physicians, overcoming the disconnect between their knowledge of PF (Table S5).

Genetic counsellors additionally stressed the importance of psychosocial support, having resources readily available, and scheduling adequate time ensuring there is adequate time to discuss complex results.

> *‘Some of the pulmonary fibrosis cases I’ve seen, from a psychosocial perspective, are some of the really complicated ones. They need support and it’s not always the physician that’s available to give that. I am someone that they can call’ (GC_2)*.A summary of overarching themes is provided in Figure 1.

**Figure 1.**
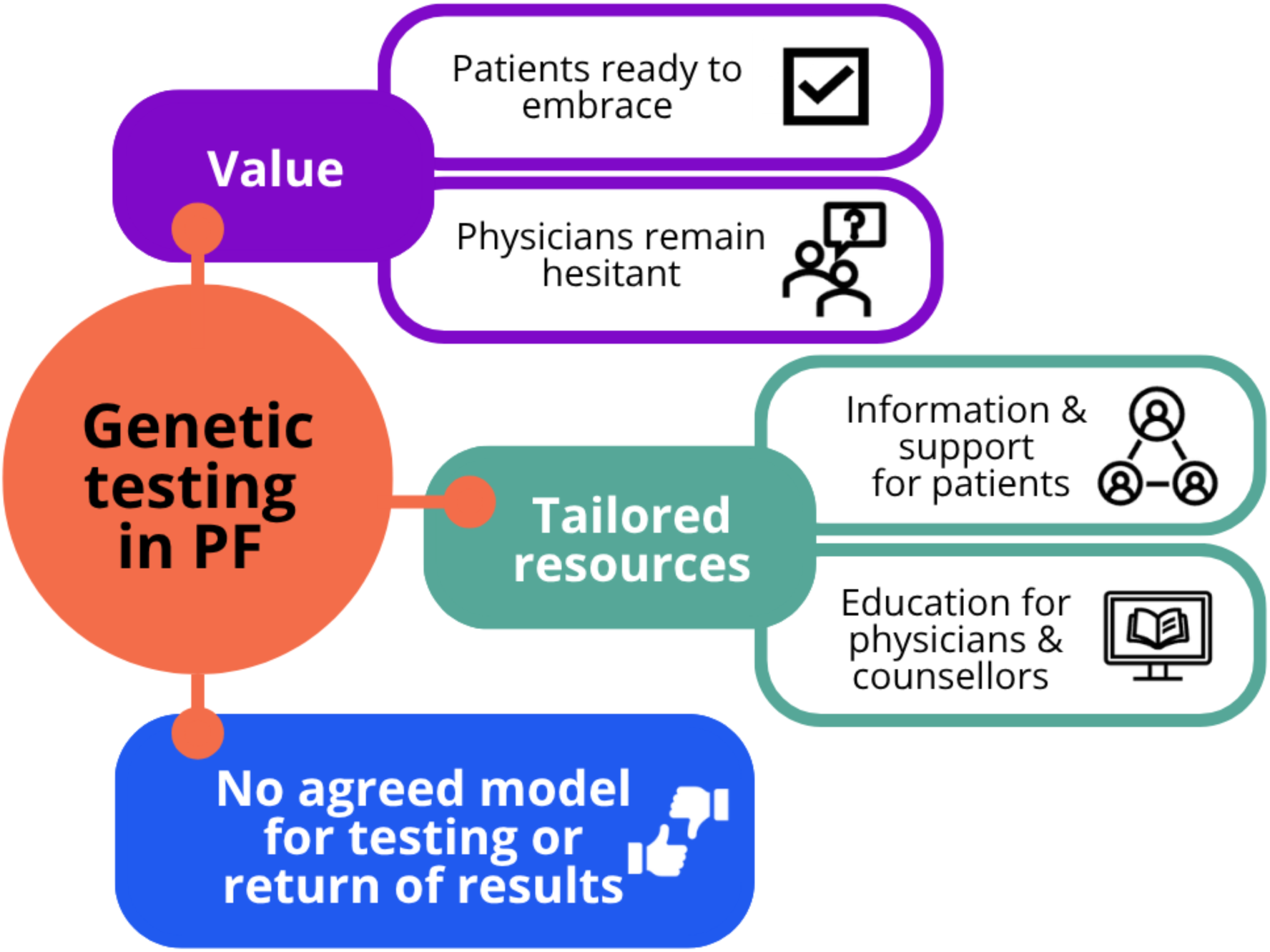
**Summary of themes**

## Discussion

Our qualitative study found genetic testing was universally valued by people living with PF and HCPs. People living with PF were ready to embrace genetic testing now, and motivated to know more about their disease. Respiratory physicians however remained hesitant due to uncertainty regarding what genetic results meant for clinical decision-making. All stakeholders desired more information, but the nature of this varied. Patients want information to understand genetic language more easily; respiratory physicians want to know how to practically carry out testing and return results; while genetic counsellors want help around understanding PF. Ultimately, there was no optimal model for returning genetic results in PF.

Whilst there are a number of genetic variants currently implicated in PF, evidence for their clinical impact is still emerging(14). Pathogenic telomere gene variants and resulting clinically significant short telomeres are major contributors to inherited PF(6, 15). Knowledge of telomere-related PF can have actionable implications for clinical practise(16), supporting clinical decisions around antifibrotic therapy, transplant timing and management(17), and provide useful prognostic information on disease progression(6, 15, 16). Lesser-known genetic variants implicated in PF lack sufficient evidence to assess pathogenicity, creating ongoing uncertainty around their usefulness(14). Similar uncertainty surrounding clinical genetic test results has been, and continues to be, faced across the spectrum of genetics. Cancer specific genetic testing guidelines, encompassing pre and post-test counselling, encompass information for patients regarding the limitations with genetic testing(18). Acknowledging testing uncertainty paradoxically supported patients to proceed with testing, and patients acknowledged the need to build greater evidence and larger registries surrounding lesser-known variants(18). A comparable trajectory for emerging gene variants in PF may similarly evolve with ongoing research and increased testing(19).

All stakeholder groups want more genetic resources and support, however different needs were evident. Genetic resources for patients with PF and their families are currently limited(8). People living with PF are uncertain how genetic results are disclosed or what the insurance implications may be(8). The lack of easily accessible resources may also contribute to the low prevalence of genetic testing in PF(6, 20). Less than 30% of respiratory physicians routinely order genetic testing, with the majority conducted in familial cases, within specialist centres(6). National and international guidelines are absent, and recommendations are inconsistent, including who to test, how to test, how to disclose results, and how at-risk relatives should be managed(21). This highlights the need for standardised pathways to improve the application of genetic testing in PF. Pulmonary fibrosis is a rare disease, and it is not surprising genetic counsellors reported a need to deepen their understanding of PF. Similarly in cancer, genetic counsellors report high confidence counselling patients with well-known cancers, however confidence is lower in lesser-known cancers(22). Genetic guidelines endorsed by the American College of Medical Genetics and Genomics (ACMG) outline the importance of clinician education and training, ensuring safe and effective application of genetic testing clinically(23). As genetic testing practises evolve in PF, adequate education and training are a necessary consideration(21).

There was no optimal model for returning genetic results in PF. While genetic counsellor and respiratory physician preferences for return of results by specialists align with current recommendations made by the Pulmonary Fibrosis Foundation Genetic testing work group(6), the European Respiratory Society(14) and more broadly the HGSA(24), this did not necessarily reflect perspectives of patients and families, who desired relational support with someone they know well. The European Society of Human Genetic (ESHG) and HGSA clinical genetic framework recommends genetic results be returned by a clinician with genetic expertise(24, 25). This model would be achievable for patients with proximity to specialist ILD centres, while those receiving care in rural or non-specialist settings may have limited access to such expertise. Similar inequities have been faced and continue to challenge genetic services across many diseases, such as cancer(18). As genetic testing becomes increasingly critical for diagnosis and management of lung disease, more accessible care models will be necessary.

Study strengths included participants from a broad range of stakeholder groups located across Australia, including patients and HCPs. Health professional perspectives encompassed a range of disciplines involved in care of PF and/or genetic testing, revealing diverse perspectives and information needs. Our study does have several limitations. Most physicians interviewed worked in metropolitan centres (91%), where the awareness of genetic testing was high. In addition, the recruitment of people living with PF was conducted using convenience sampling. Participants who were from non-English-speaking backgrounds were not interviewed, therefore their perspectives remain unknown. The low caregiver sample limited the interpretability of caregiver specific findings, future studies should aim to recruit larger caregiver cohorts to better understand caregiver needs and perspectives.

## Conclusion

Genetic testing is valued by people with PF and their HCP. Whilst people with PF want to embrace genetic testing now, uncertainties regarding its clinical implication cause hesitancy for physicians, and there is not agreed model for return of results. Tailored education and resources for patients, physicians and genetic counsellors will be required to support genetic testing into the future for PF.

## Data Availability

All data produced in the present study are available upon reasonable request to the authors

## Data Availability

All data produced in the present study are available upon reasonable request to the authors

## Acknowledgements

Grateful to the people living with PF and health professional for sharing their experiences and the Breathing Easier Together Advisory Group for their assistance.

## Conflicts of interest

SR- nil to declare

NSC - nil to declare

JD - nil to declare

CSW - nil to declare

AEH - nil to declare

The authors report no commercial support for the work presented in this manuscript. The authors have no commercial relationships in the past three years that could be perceived as relevant to the submitted work. The authors report no non–financial associations relevant to the submitted manuscript.

## Author contributions

Concept and design: SR, NSC, JD, AEH Data acquisition: SR

Data analysis and interpretation: SR, CSW Manuscript drafting: SR, NSC, CSW, JD, AEH

Critical review of manuscript and approval of final version: SR, NSC, CSW, JD AEH

