## Supplemental files for "Exploring the views of people living with pulmonary fibrosis and health professionals on genetic testing in PF: A qualitative study"

Exploring the views of people living with pulmonary fibrosis and health professionals on genetic testing in PF: A qualitative study 
S Rawlings, NS Cox, CS Wan, J Dickinson, AE Holland


Supplementary Material


Interview guide
Table S1 People living with PF interview guide. 
Area of Interest 
Descriptive Question 
Experience of testing 
 
 
Can you tell me about you and/or anyone from your family that may have experience with genetic testing in relation to PF? 
Probing questions:
If genetic testing WAS offered: 
• What is your perception of genetic testing? 
• Can you describe for me how it was offered?  
• If you were offered genetic testing again in the future, would you consider it? 

If genetic testing was NOT offered, can you tell me about your thoughts on genetic testing in relation to PF. 
• What is your perception of genetic testing? 
• If you were offered genetic testing in the future, would you consider it? 
Return of results 
Following genetic testing you may have received results. 
How have genetic results been returned to you?
Or  
In an ideal situation, how do you think your genetic results could best returned to you?  
Probs:
• Who do you think should tell you about your results?
• What type of result do you expect to receive back? (
• What support would you need if you were to undergo genetic testing? 
Can you tell me about any information or resources that you found or feel would be an important as part of this process? 
Tell us about any support that was provided to you before, during and after receiving your genetic results? 
Describe how you would feel if you didn’t get a clear answer?  
Returning results: Genetic testing variants
 

Information: Genetic variants are normal. We all have genetic variation, and largely these variants don’t impact our health, instead just contribute to why we look different to others (different hair colour, eyes or heights etc.). Some variants, however, do contribute to disease, either by causing or increasing our risk of developing it. When we are “uncertain” about a particular variant it means we don’t know if that variant is related to their disease, or if the variant is just part of what makes them, them (because they are an individual).  
 
Genetic tests may find a genetic result that we know might cause PF, but sometimes they also identify a genetic finding where we are uncertain if its disease causing or not.  Sometimes these results turn out to be important after more research, but sometimes they don’t.  
 
Can you describe to me which genetic results you feel should be returned to you and why? 
Future: Genetic testing in research  
 

Currently in Australia, and internationally, there are opportunities for PF patients to contribute to research. This research may include genetic testing of patients and their family members. 
 
Can you tell me your perspective about the role of genetic testing in research for people with PF? 
Genetic testing in clinical trials 
In the future, it is possible that there will be treatments for people with PF that are based on their individual genetic cause of their disease. To move towards this goal, we are now starting to see clinical trials that target specific genetic results.  To participate in such a trial, a person with PF would have to have a genetic test to see if they were suitable. 
 
If you were approached about such a trial, how would you feel about undergoing a genetic test to determine your eligibility? 
Close of interview 
 
What would you suggest to health professionals about the best way to return genetic results to a person with PF? 
Is there anything else you would like to say or discuss today that we have not yet talked about in this interview? 


Table S2 Health professional interview guide
Area of Interest 
Descriptive Question 
Experience of testing & current practise. 
 
 
 
Tell me about your perspective around the role of genetic testing for people with PF? 
Tell me about your experience with genetic testing? 
Probe: Describe what is or has been your role within this space? (offering, return of results, support/ counselling, ongoing health care) 
If you have offered genetic testing, can you describe for me in more detail the process by which you offered the patient genetic testing?  
Can you tell me about anything that has prevented you from offering genetic testing to your patients? What would be required for you to offer genetic testing to your patients? 
Returning results
Can you describe your experience with returning genetic results? 
Probes:
• What results were returned back to the patient? 
• When are results typically returned? 
• Who was in charge of returning these results? 
• When returning genetic results what makes you feel confident to deliver these results?  
• What information and/or training was provided to you to return these results? 
• Are there any resources/support you have accessed/found useful to deliver the return of results? Is there any additional information or education you feel would be important to have as part of this process?   
• Describe any aspects that you found not so useful or negative when you returned these results? 
• Describe any aspects that you found useful or positive when you returned these results? 
Returning results: genetic variants
Genetic tests might find genetic variants that we know are disease causing in PF, but sometimes they also find genetic variants where we don’t know whether they matter or not. Sometimes these variants turn out to be important after more research, but sometimes they don’t.  
 
Can you describe to me what genetic results you feel should be returned and why?  
If a colleague asked you about delivering genetic results clinically, what advice or recommendations would you offer them? 
Ideal Framework for genetic testing  
 
 
In your opinion can you describe for me the ideal model in which genetic results are returned? 
Does this differ to what you currently do? 
Probes:
• Who should provide feedback on results (HCP, clinical trialist etc) 
• When should results be ideally returned? 
• What is the best mode to deliver and return results? 
• When is the most ideal time to involve the family in the return of results?
Future: Genetic testing in research  
 
In the future, it’s possible that there will be treatments for people with PF that are based on their individual genetics. To move towards this goal, we are now seeing clinical trials that target specific genetic variants.  
In order to participate in such a trial, a person with PF would have to have a genetic test to see if they were suitable. 
 
Can you briefly tell me about your experience of genetic testing for research purposes?  
Do you have any experience around genetic testing and/or the return of genetic results for research? 
Can you describe your current attitudes and feelings towards the return of genetic results in genome studies/ research? 
Close of interview 
Is there anything else you would like to say or discuss today that we have not yet talked about in this interview? 


Main themes 
Table S3. Theme 1 Genetic testing is valued. 
Subtheme
Supporting Quote
Participant ID
People with PF and HCP recognise the value of genetic testing, but ongoing barriers exist. 
I think it’s an excellent idea. I really do. I’d be interested to know if there’s any link, with family history. 
P1

The fact that it took this sort of process for me to be diagnosed, whereas if it was possible to look at other ways of diagnostic testing, particularly if it’s looking at within families or something like that, then I don’t have a problem
P4

I see that genetics just adds to the story about what are the chances of this patient getting progressive disease, responding to anti-fibrotic treatment. Anything we can do to chip away and make things a little bit easier you'll get a lot of buy in from clinicians, I think.
RP_9

What's becoming clear over the last 10 years really is the importance of genetic testing in ILD. The more that I learn about it the more it’s clear to me that how important it is.
RP_5

The process from referral (to geneticist) to results on average is 18 months to 24 months.
RP_3

It's lack of access, easy access to genetic testing. It's just not widely available. It's very difficult to do from regional and remote areas in particular. 
RP_4

We all recognise the importance of genetic testing but it’s a big thing to add on to what we’re already doing. It’s an unmet need, but we don't have the resources for it either at this point. 
RP_6

We need more access to genetic testing and geneticists and genetic counsellors in order to have the right framework for testing. It’s having it done in the right framework and having the right people around to help support the clinician.
RP_5

Many institutions don't have a clear pathway for mainstream genetic testing.
RP_9

The results take about six months, which is unacceptable. 
Gen_2
People with PF are ready to embrace testing now.
The more information the better because its information that can assist you to deal with it. The more aware you are, the better.
P18

Genetic testing is part and parcel...I might be the first in my family line, who knows, at this point this is why you are trying to find out.
P15

Any knowledge from her disease cannot be bad; it must be an advantage to have.
C3

It’s really not going to change my outcome, but it could potentially change other people’s outcomes and that’s what I’m more interested in. I mean, new treatments, hopefully they come in every day, and I guess that’s what we live in hope of.
P8
Respiratory physicians remain hesitant, driven by uncertain implications for clinical practise. 

I would need evidence that it makes a difference. At the moment we have a horrible disease with crap drugs, and they all continue to get worse invariably. Someone would need to tell me that doing a gene test is going to be helpful in the future. I think if there’s proven benefit, absolutely, we’ll do anything. 
RP_10

We should have genetic testing in our armoury of what we can offer patients. I think there’s a few questions on how it translates to patient care, it is not clear.
RP_06

(Variants of uncertain significance) That's part of the reason why we don't do routine genetic testing. Because we don't really know what to do with them or what their significance is. And there's no difference in how you treat the patients for it. 
RP_1

There is a lot of genetic uncertainty that we just don’t know or don’t have tests that support that testing. 
RP_13

When we order a test, we’ve got to have an idea about how we’re going to act on those results. Until I understand what the test means then I wouldn't really be comfortable in ordering it.
RP_12

For (genetic testing) to become part of our routine management I suppose there would have to be more clinical data.
RP_9


Table S4: Theme 2 More information and support are desired, but needs differ.
Subtheme
Supporting Quote
Participant ID
Genetic knowledge is variable across people with PF, caregivers and respiratory physicians, other professionals lack PF disease clarity
It’s beyond my scope of understanding. Current resources are not fit for purpose
P18

It just sounds so complicated. There is an overwhelming amount of information, when you’re at a 15-minute medical interview there’s so much to take in, that often you don’t take it all in, or you don’t understand everything.
P12

She did provide me with a sheet of information, but it does seem beyond my scope of understanding.
P4

They were good at providing what information they had but it does seem beyond my scope of understanding. Maybe understanding the objective of it a little but more.
P18

I don't have all the clinical knowledge and a lot of the questions and things that come up from patients is the clinical side of things.
GC_2

I’ve been asked into clinic when I’ve had a significant role, some of the clinicians have asked me if I wanted to be part of that return but I always feel a bit uncomfortable because I’m not a medical doctor or a counsellor.
Gen_1

I think it's part of that clinical care that we should be offering it, but we've got to find who, when, why and how.
RP_6

I see is that geneticists, most of them bar one have no idea about pulmonary fibrosis.
RP_3
People with PF want resources and accessible support to understand genetic language. 


I would like to be clear what exactly is going to happen. I think it’s important that you have got the one -to one dialogue, because I might have entirely different questions to somebody else.
P4

I guess I'd probably like to have information from other patients who have experienced (genetic testing). 
P12

I'd like to see information in print and as a video or something like that as well, something that I can study and try and understand.
P12

You're an adult and you need to be able to make your own decisions, and you can only make that decision if you know what the hell's going on.
P9

If I have a question, I could wait a month, two months, three months before I can get an appointment.
P1

Like with any kind of support access is a thing….being able to pick up the phone and call someone, send a text, and getting a response.
P10

Clarity of the (testing) procedure, just talking you through. 
P19

Current resources are not fit for purpose. I think there’s a side of genetics that seems a bit more complex to me.
P18

It’s frustrating not to be clear, but as long as you have the discussion about what could happen, what could not happen, how to go about managing whatever you manage to find out, that’s all you can expect. 
P1
Health professionals want training, pathways, and clinical support.

As long as there is a clear and accessible pathway so that we can make a referral in a very timely fashion and get them seen, that’s probably the most important element
RP_11

The whole thing about this whole space is that there needs to be education, upskilling with education about the meaning of these tests.
RP_8

A referral from, where to send blood as a starting point. Also a bit of education, at the moment everything is a little ad hoc, a little bit of discussion here and there.
RP_9

It's more a matter of understanding what it means and that requires skill and counselling
RP_8

I guess it is interpretation of the tests, you know, what does it mean for their children, for their siblings, other family members because I would need more training to be able to help the patients more
RP_2

There’s lots of things that you learn about by working as a team, but I think if you could put that into a resource, that would be excellent.
GC_1

You can make decisions as a group which is much better because before it was us and one person.
GC_1

It’s a very close relationship between the genetic counsellor and the clinician.
Gen_1

A combined approach is usually beneficial for the patient, but also for the clinicians.
Gen_2


Table S5: Theme 3 There is no optimal model for returning genetic results in PF.
Subtheme
Supporting Quote 
Participant ID
Views are varied on who is best to return genetic results.
The medical person that I have a lot of faith in, and that I have a good rapport with I guess 
P14

I would expect my specialist would tell me about it and discuss it with me at the time.
P14

Probably my GP I have known for a very long time’
P13

I would expect my specialist would tell me about it’
P14

She wouldn’t give dame who gave her the results. I guess the current doctor she’s under at the moment would be the one. But really I don’t think it matters.
CO3

Well obviously, somebody with some expertise.
P17

Anyone, to tell you the truth. I wouldn’t be fussy
P8

I think the person that you initially have the contact with that pertains to that area because they’re the people that are the most familial with it and certainly the respiratory doctors and geneticists
P18

I would think that it would sit with the tertiary hospitals. It won't be a primary care thing. It has to sit with a respiratory physician as well.
RP_1

The results that I have initiate myself where I have done the consenting and testing though. I would give the results back.
RP_7

It should be always delivered by a clinician expert in those decisions 
RP_2

It’s unfair to ask someone who doesn’t know about it to return the result in an accurate fashion. I try not to return the results because I think it’s much better done by a geneticist or a genetic counsellor 
RP_5

The clinician that orders the test gives the results and does the follow up plan for the positives or for the uncertain one
Gen_4

Negative results can be delivered by respiratory physicians
RP_1

I think the preliminary result from the genetics with a follow-up discussion with respiratory would be appropriate
RP_6

I think that the return of results really would work best for something like pulmonary fibrosis in a multidisciplinary team
Gen_2

Because of the expertise of the physicians, I think they absolutely can deliver the results, but they don't have the time nor necessarily the understanding of all the different pathways of how to deal with all of the implications that come with doing genetic testing.
GC_2
Logistics to support return of genetic result include resources and multidisciplinary collaboration.


I think that face-to-face with the specialist who initiated it is very important.
P7

I am happy for the information to come vis email, but it’s always helpful then, if there are any sort of out of whack results, it’s always helpful to follow that up with a discussion with a medico of some kind.
P1

I mean as a patient, it’s very comforting to know that, well it's not just one guy dealing, but he actually does talk to others.
P9

I would like to have the information soon as possible. 
P12

I think a personal explanation is the way to go to make sure that people aren’t left to try and interpret medical data 
P16

I think it can’t always be a one size fits all 
P4

I still didn’t like doing everything on the computer.  I know there’s a lot of benefits of using the computer and you save paper and, you know, and you can save everything.  It doesn’t have to take up space in your office and all that sort of stuff.  But it’s just not the same as real life – in real life.
P14

I think a team should communicate, and I know it’s difficult and expensive, but I would say that it’s more than just one meeting
P10

Counselling needs to be in place to address the results and what implies the results.
RP_8

You really need someone to do the counselling for family. Because I think it will be very hard to squeeze in counselling for everyone by the respiratory physicians with their family as well
RP_1

Having good links with the laboratory so that you can have a dialogue about how likely they think it might be pathogenic, those things are really important. 
Gen_2

You need to talk about it. It's not good enough for a person just - a clinician to manage a super complicated patient just by themselves in private, it's just not good enough, like medicine is beyond - you need multidisciplinary expert care and discussion.
RP_9

It would be nice to have a handout I can give to the patients because it's really not something that can be easily understandable and it would be hard for them to find information themselves.
RP_1

(Supporting family understanding) if there was some sort of, like, printed booklet or something they could read themselves after I’d explained it to them
RP_14

I think that returning results really would work best for something like pulmonary fibrosis in a multidisciplinary team.
Gen_2

Genetic medicine is definitely best practiced in teams.
Gen_3

It would be very important to provide them written information and other resources they can access to understand it from an expert group.
RP_12

This is a patient group that’s over 50. Mostly over 60. They don’t want to go onto a website. They don’t want something amorphous. They want a piece of paper
N_2

That does require time explaining those concepts, but it doesn't have to be done by genetics.
Gen_4

It’s very much guided by the patient. Sometimes people want to have their family involved from the very beginning, sometimes they want to know what – the answer first before they involve their family.
RP_5

If you have established that relationship already, then I don’t think there is a big issue with delivering the result by telehealth
RP_11
